# High incidence of sotrovimab resistance and viral persistence after treatment of immunocompromised patients infected with the SARS-CoV-2 Omicron variant

**DOI:** 10.1101/2022.04.06.22273503

**Authors:** Sammy Huygens, Bas Oude Munnink, Arvind Gharbharan, Marion Koopmans, Bart Rijnders

**Affiliations:** Department of Internal Medicine, Section of Infectious Diseases and Department of Medical Microbiology and Infectious Diseases, Erasmus MC, University Medical Center, Rotterdam, The Netherlands; Department of Viroscience, Erasmus MC, University Medical Center, Rotterdam, The Netherlands

## Abstract

**Background:** Sotrovimab is a monoclonal antibody that neutralizes SARS-CoV-2 by binding to a highly conserved epitope in the receptor binding domain. It retains activity against the Omicron BA.1 variant and is used to treat immunocompromised patients as they are at increased risk for a severe outcome of COVID-19.

**Methods:** We studied viral evolution in 47 immunocompromised patients infected with Omicron BA.1 or 2 and treated with sotrovimab. SARS-CoV-2 PCR was performed at baseline and weekly thereafter until Ct-value was ≥ 30. All RNA samples were sequenced to determine the variant and occurrence of mutations, in particular in the Spike protein, after treatment.

**Results:** Twenty-four (51%) of the 47 patients were male and their median age was 63 years. Thirty-one (66%) had undergone a solid organ transplantation and 13 (28%) had received prior B-cell depleting therapy. Despite a history of vaccination, 24 of 30 patients with available data on anti-SARS-CoV-2 IgG Spike antibodies prior to treatment with sotrovimab had very low or no antibodies. Median time to viral clearance (Ct-value ≥ 30) after treatment was 15 days (IQR 7-22). However, viral RNA with low Ct-values was continuously detected for at least 28 days after treatment in four patients infected with BA.1. Mutations in the Spike protein at position 337 or 340 were observed in all four patients. Similar mutations were also found after treatment of two patients with a BA.2 infection but both cleared the virus within two weeks. Thus following treatment with sotrovimab, spike mutations associated with reduced in vitro susceptibility were detected in 6 of 47 (13%) patients.

**Conclusion:** Viral evolution towards resistance against sotrovimab can explain treatment failure in most immunocompromised patients and these patients can remain infectious after treatment. Therefore, documenting viral clearance after treatment is recommended to avoid that these patients unintentionally become a source of new, sotrovimab resistant, variants. Research on direct acting antivirals and possibly combination therapy for the treatment of COVID-19 in immunocompromised patients is needed.

## Introduction

Sotrovimab is a monoclonal antibody that neutralizes SARS-CoV-2 by binding to a highly conserved epitope in the receptor binding domain of sarbecoviruses. It is one of the few approved monoclonals that retains activity against the Omicron BA.1 variant of concern (VOC).^1^ Immunocompromised patients are at increased risk for a severe outcome of COVID-19 even after full vaccination against SARS-CoV-2. Indeed, the vaccination response is often reduced and can be completely absent in patients with combined T-cell and B-cell dysfunction.^2,3^ Furthermore, prolonged viral replication and evolution has been described in the immunocompromised host.^4,5^ Given the risk for a poor outcome, monoclonal antibody-based therapy is used to treat these patients. Especially during antibody *mono*therapy, viral evolution towards resistance against the antibody can be expected when replication is not sufficiently contained. Recently, the selection of mutations in the spike protein of the Delta VOC in 4 of 100 patients treated with sotrovimab was reported and all 4 were immunocompromised.^6^ Specifically, mutations at position 337 and 340 known to reduce susceptibility to sotrovimab were found.^7^ We studied viral evolution in 47 immunocompromised patients treated with sotrovimab for an infection with the Omicron VOC.

## Methods

At Erasmus University Medical Center in Rotterdam, sotrovimab became available on January26^th^,2022. It is used to treat immunocompromised patients infected with the SARS-CoV-2 Omicron VOC in the outpatient and inpatient setting. A SARS-CoV-2 PCR was performed at baseline and weekly thereafter, also after discharge until the PCR Ct-value was ≥30.^8^ Baseline and follow-up samples with a Ct-value <30 were sequenced on the Nanopore platform.^9^ Successful sequencing was defined as at least 90% of the genome covered with at least 30x coverage. The study was approved by the institutional review board under number METC2021-0309.

## Results

Of the 47 patients treated, 24(51%) were male, the median age was 63(IQR 51 – 67), 31(66%) had undergone an organ transplantation, 13(28%) received B-cell depleting therapy and 5 treated with other immunosuppressives for auto-immune disease. 31 of 47(66%) patients were hospitalized when sotrovimab was given. In 30 patients, an IgG Spike antibody titer(Liaison IgG Trimeric S, Diasorin) was measured in the 30 days preceding sotrovimab therapy. Despite a history of at least 2 mRNA vaccinations in all and 3 or more in 24, Spike antibodies were negative in 18 and very low(<300 BAU/ml) in 25.

The median time to a Ct-value ≥30 was 15 days(IQR 7-22 days; range 3-36 days) but in 4 patients low PCR Ct-values persisted 29, 36, 49 and 49 days after treatment(Figure 1). Sequencing of at least one RNA sample was successful in 32 patients(25 BA.1 and 7 BA.2) and for 17 patients sequencing of a baseline as well as follow-up RNA samples was successful. In 6 of these 17 patients(35%), key spike mutations on positions 337 and 340 known to confer in vitro resistance to sotrovimab were found(Figure 1). This was the case for 4 of 25(16%) BA.1 infected patients and for 2 of the 7(29%) infected with BA.2. In a third BA.2 infected patient, a D796Y mutation was found but its impact on the sotrovimab IC50 is unknown. In all 4 patients with prolonged viral replication and BA.1 infection, mutations at position 337 or 340 of the Spike protein were found.

**Figure 1:**
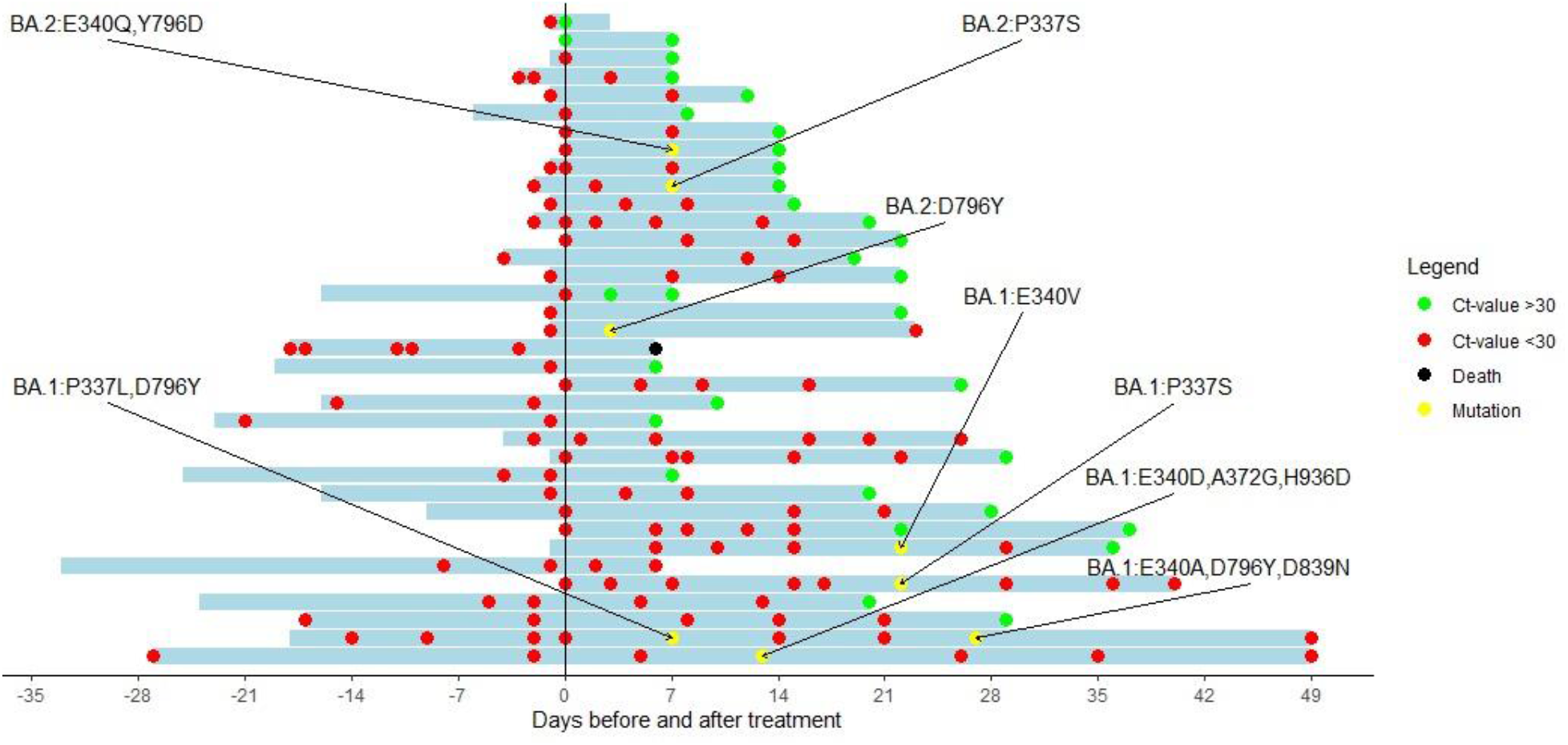
Cyclic threshold value of patients treated with sotrovimab with a baseline RT-PCR and at least one follow-up RT-PCR available. The y-axis indicates the day of sotrovimab infusion. Red dots represent a Ct-value <30. Green dots represent a Ct-value of >30. Black dots represent the dead of a patient. Yellow dots represent a sequence showing new Spike mutations compared with the baseline Sequence.

## Discussion

Following treatment with sotrovimab, spike mutations associated with reduced in vitro susceptibility were detected in 6 of 47 patients.^7^ Furthermore, more than 4 weeks after treatment with sotrovimab, 4 of them continued to have a high viral load. Despite the extremely low IC50 concentrations of sotrovimab against BA.1, genotypic resistance was detected after treatment in 4 of 25(16%) of the BA.1 infected patients. The high frequency of mutations found after treatment of patients with BA.2(43%) may be caused by the already 16-fold increased IC50 of sotrovimab against BA.2.^10^

Our observations show that viral evolution towards resistance can explain treatment failure in the majority of patients. Immunocompromised patients unable to clear SARS-CoV-2 despite antiviral therapy can serve as a source of new variants and should be closely followed until viral clearance is documented. Research is urgently needed to evaluate the value of direct acting antivirals in this patient group. Similar to the treatment of HIV, combination therapy may be required to reduce the risk for resistance.

## Data Availability

All data produced in the present study are available upon reasonable request to the authors

## Notes

### Competing Interest Statement

The authors have declared no competing interest.

### Funding Statement

This work has received funding from the European Unions Horizon 2020 research and innovation programme under grant agreement numbers 874735 (VEO) and 101003589 (RECoVER) and from ZonMW under grant 10150062010005

### Author Declarations

Ethics committee/IRB of Erasmus Medical Center (Rotterdam)gave ethical approval for this work

## References

1. VanBlargan LA, Errico JM, Halfmann PJ, et al. An infectious SARS-CoV-2 B.1.1.529 Omicron virus escapes neutralization by therapeutic monoclonal antibodies. Nature Medicine 2022; 28(3): 490–5.

2. Haggenburg S, Lissenberg-Witte BI, van Binnendijk RS, et al. Quantitative analysis of mRNA-1273 COVID-19 vaccination response in immunocompromised adult hematology patients. Blood Adv 2022; 6(5): 1537–46.

3. Sanders JF, Bemelman FJ, Messchendorp AL, et al. The RECOVAC Immune-response Study: The Immunogenicity, Tolerability, and Safety of COVID-19 Vaccination in Patients With Chronic Kidney Disease, on Dialysis, or Living With a Kidney Transplant. Transplantation 2021; 106(4): 821–34.

4. Corey L, Beyrer C, Cohen MS, Michael NL, Bedford T, Rolland M. SARS-CoV-2 Variants in Patients with Immunosuppression. N Engl J Med 2021; 385(6): 562–6.

5. Harari S, Tahor M, Rutsinsky N, et al. Drivers of adaptive evolution during chronic SARS-CoV-2 infections. medRxiv 2022: 2022.02.17.22270829.

6. Rockett R, Basile K, Maddocks S, et al. Resistance Mutations in SARS-CoV-2 Delta Variant after Sotrovimab Use. New England Journal of Medicine 2022.

7. Cathcart AL, Havenar-Daughton C, Lempp FA, et al. The dual function monoclonal antibodies VIR-7831 and VIR-7832 demonstrate potent in vitro and in vivo activity against SARS-CoV-2. bioRxiv 2022: 2021.03.09.434607.

8. van Kampen JJA, van de Vijver DAMC, Fraaij PLA, et al. Duration and key determinants of infectious virus shedding in hospitalized patients with coronavirus disease-2019 (COVID-19). Nature Communications 2021; 12(1): 267.

9. Oude Munnink BB, Nieuwenhuijse DF, Stein M, et al. Rapid SARS-CoV-2 whole-genome sequencing and analysis for informed public health decision-making in the Netherlands. Nature Medicine 2020; 26(9): 1405–10.

10. GlaxoSmithKline. Sotrovimab [package insert]. U.S. Food and Drug Administration website. https://www.fda.gov/media/149534/download. Revised March 2022. Accessed April 4, 2022.

